# Lipedema-like Phenotype and Cancer Prevalence in US Women: A Cross-Sectional Analysis of NHANES 2011–2014

**DOI:** 10.64898/2025.12.02.25341445

**Authors:** Alexandre C. M. Amato, Juliana L. S. Amato, Daniel A. Benitti

## Abstract

**Objective:** To examine the association between lipedema-like peripheral fat distribution and cancer prevalence in a nationally representative sample of US women.

**Methods:** We analyzed cross-sectional data from 2,818 women aged 20 to 59 years in NHANES 2011–2014. The exposure was leg-to-trunk fat ratio (LTR) measured by DXA. Multivariable logistic regression estimated odds ratios (OR) for cancer history, adjusting for BMI, race/ethnicity, smoking, and education. Primary models excluded age to estimate the phenotype’s total effect (viewing age as a mediator).

**Results:** Cancer prevalence was 4.6%. Women with cancer exhibited lower LTR compared to controls (*P*=.0048). Each 1-SD increase in LTR was associated with 20% lower cancer odds (OR 0.80; 95% CI 0.67–0.95; *P*=.011). This inverse association was robust among women without obesity (OR 0.67; 95% CI 0.53–0.85; *P*=.0007) and showed a consistent trend among women with obesity (OR 0.74; *P*=.07). Descriptive analyses confirmed that women with cancer presented lower LTR within all pre-menopausal age decades.

**Conclusions:** A peripheral fat distribution consistent with the lipedema-like phenotype is inversely associated with cancer prevalence. This protection appears robust in women without obesity and follows a similar directional trend in women with obesity, suggesting that gluteofemoral adipose tissue may serve as a protective immunometabolic buffer.

**Clinical Trial Registration:** Not applicable (Analysis of publicly available NHANES data).

## INTRODUCTION

Lipedema is considered a chronic adipose tissue disorder characterized by symmetrical, disproportionate accumulation of subcutaneous fat in the lower extremities, typically sparing the hands and feet, and accompanied by pain, tenderness, easy bruising, and orthostatic edema (1). First described by Allen and Hines at the Mayo Clinic in 1940 (2), the condition affects almost exclusively women and is frequently misdiagnosed as general obesity, lymphedema, or venous disease. Although population-based data remain limited, an estimated prevalence of 12.3% among adult women, is a substantial public health concern rather than a rare condition (3). Diagnosis remains essentially clinical, based on the pattern of fat distribution and characteristic symptoms, which motivated the development and validation of screening tools achieving over 90% accuracy in predicting affected individuals (4, 5). Recent consensus guidelines have reinforced that lipedema cannot be adequately captured by body mass index alone and that more specific anthropometric and body composition measures are needed to characterize its distinctive peripheral fat phenotype (6).

Beyond regional adipose accumulation, converging evidence indicates that lipedema is a systemic disorder with immune, microvascular, and metabolic abnormalities distinct from those observed in common obesity (7). Histologic and molecular studies have described adipocyte hypertrophy, microvascular dilation, macrophage infiltration, and altered cytokine profiles in affected tissue, supporting a model of chronic low-grade inflammation. Building on this framework, recent investigations have demonstrated a paradoxical immunoglobulin G food reactivity signature in women with lipedema, characterized by more frequent positive food-specific IgG tests despite markedly lower total IgG levels, consistent with mucosal immune dysregulation and impaired barrier function (7). Complementary genetic studies have found a high prevalence of HLA-DQ2 and HLA-DQ8 risk haplotypes in symptomatic patients, with clinical improvement reported following gluten-free dietary interventions (8). Together, these findings suggest that the distinctive peripheral adipose phenotype in lipedema is embedded in a broader immunometabolic context rather than representing a purely mechanical fat storage disorder. However, this distribution is rarely discussed in oncology.

Obesity is an established risk factor for at least 13 malignancy types, with mechanisms including chronic inflammation, insulin resistance, altered adipokine secretion, and sex hormone dysregulation. However, the relationship between body fat distribution and cancer risk appears heterogeneous. Central and visceral adiposity, reflected by increased trunk fat mass and waist circumference, has been consistently associated with higher cancer incidence and mortality, whereas gluteofemoral and peripheral subcutaneous fat may exert a neutral or even protective role. Peripheral adipose tissue may act as a “metabolic sink” that buffers lipotoxicity and systemic inflammation through preferential fatty acid uptake and adiponectin secretion. In this framework, the lipedema-like phenotype can be viewed as an extreme form of peripheral adiposity in the lower body, often coexisting with elevated body mass index but with distinct microvascular and immune features. Despite this conceptual plausibility, the association between a lipedema-like pattern of fat distribution and cancer has not been examined in large, population-based datasets with standardized body composition measurements.

We therefore hypothesized that a lipedema-like peripheral fat distribution, quantified by the ratio of leg to trunk fat mass on dual-energy x-ray absorptiometry, would be associated with lower cancer prevalence in women, particularly among those with obesity, independent of overall adiposity (9, 10). Using nationally representative data from the National Health and Nutrition Examination Survey (NHANES) 2011–2014, we sought to characterize the dose-response relationship between the leg-to-trunk fat ratio and cancer prevalence, to evaluate whether an extreme peripheral distribution consistent with the lipedema-like phenotype confers relative protection, and to compare the predictive performance of this anthropometric marker with traditional indices such as body mass index and trunk fat mass.

## METHODS

### Study Design and Population

We conducted a cross-sectional analysis of data from the 2011–2012 and 2013–2014 cycles of the NHANES, a nationally representative study of the noninstitutionalized US civilian population. Women aged 20 to 59 years with complete whole-body dual-energy x-ray absorptiometry (DXA) data and cancer history were eligible for inclusion. The upper age limit was dictated by the NHANES body composition examination protocol, which restricted whole-body DXA scans to participants aged 8 to 59 years during these cycles. After excluding participants with missing body composition measurements, incomplete covariate data, or current pregnancy, the final analytic sample comprised 2,818 women (Figure S1). This study followed the Strengthening the Reporting of Observational Studies in Epidemiology (STROBE) reporting guidelines (Supplementary File 1)

### Variables and Measurements

The exposure of interest was peripheral fat distribution, quantified by the leg-to-trunk fat mass ratio derived from DXA scans performed using Hologic Discovery A densitometers following standardized protocols. This ratio serves as an objective proxy for the lipedema phenotype, characterized by disproportionate adipose accumulation in the lower extremities relative to the trunk. For primary analyses, the leg-to-trunk ratio was modeled as a standardized continuous variable (z-score) to estimate odds ratios per 1–standard deviation increment. For dose-response assessment, participants were classified into quartiles, with the highest quartile representing the most peripheral, lipedema-like fat distribution pattern.

The primary outcome was self-reported history of any malignancy (MCQ220), with cancer type further ascertained via questionnaires MCQ230A-C. Given the cross-sectional design of NHANES, this outcome represents cancer prevalence or survivorship rather than incidence. Covariates included body mass index (BMI), race/ethnicity (categorized as Mexican American, Other Hispanic, Non-Hispanic White, Non-Hispanic Black, and Other/Multi-racial), smoking status (never, former, current), and education level.

### Ethical Approval

The NHANES protocol was approved by the National Center for Health Statistics Research Ethics Review Board, and all participants provided written informed consent.

### Statistical Analysis

All analyses incorporated examination weights rescaled to the combined sample size to account for the complex survey design. Descriptive characteristics were compared between women with and without cancer using design-adjusted t-tests for continuous variables and chi-square tests for categorical variables.

Multivariable logistic regression models (Generalized Linear Models with binomial family) were used to estimate odds ratios (OR) and 95% confidence intervals (CI). The primary model adjusted for BMI, race/ethnicity, smoking status, and educational attainment—confounders selected *a priori* based on established associations with adipose distribution and cancer risk.

Crucially, our analytical framework utilized Directed Acyclic Graph (DAG) theory to distinguish confounders from mediators. Age was conceptualized as a mediator because aging drives both the redistribution of adipose tissue (lowering LTR) and carcinogenesis. Consequently, primary models deliberately excluded age to capture the *total effect* of the phenotype and avoid over-adjustment bias. To estimate the *direct effect* independent of age-related pathways, we performed sensitivity analyses that included age in the model. We further evaluated the discriminative capacity of anthropometric indices by comparing the Area Under the Receiver Operating Characteristic Curve (AUC) of age-only models versus models containing age plus the specific index, using the DeLong method and likelihood ratio tests.

We assessed effect modification by obesity status (BMI ≥30 vs <30 kg/m²) and age (<50 vs ≥50 years) using interaction terms and likelihood ratio tests comparing nested models. The linear trend across quartiles was evaluated using the Cochran–Armitage test. To assess robustness to unmeasured confounding, we calculated E-values. Statistical significance was defined as a two-sided P-value < .05.

Menopausal status was defined using reproductive health questionnaire data. Women were classified as postmenopausal if they reported no menstrual periods in the past 12 months (variable RHQ031) or a history of hysterectomy (variable RHD280). For participants with missing questionnaire data, age ≥50 years served as a proxy.

All statistical analyses were performed using Python version 3.12.2, employing Pandas (v2.2.0) and NumPy (v1.26.4) for data management, along with SciPy (v1.13.1) and Statsmodels (v0.14.2) for statistical testing and regression modeling. The NHANES protocol was approved by the National Center for Health Statistics Research Ethics Review Board, and all participants provided written informed consent.

## RESULTS

### Population Characteristics and Body Composition

Among 2,818 women included in the analysis (mean [SD] age, 39.5 [11.4] years; BMI, 28.9 [7.3] kg/m²), 130 (4.6%) reported a prior cancer diagnosis. The sample was 37.6% non-Hispanic White, 23.1% non-Hispanic Black, and 17.8% Mexican American. Obesity was present in 40.3% of participants (Table 1).

**Table 1.** Comparison of body composition and anthropometric variables between women with and without a cancer diagnosis in the US population (NHANES 2011–2014).

| Variable | No Cancer | Cancer | Difference | P-value |
| --- | --- | --- | --- | --- |
| **BMI (kg/m<sup>2</sup>)** | 28.78 ± 7.20 | 30.33 ± 8.85 | +5.4% | 0.0026 |
| **Leg fat mass (kg)** | 11.58 ± 4.31 | 12.07 ± 5.11 | +4.2% | 0.0115 |
| **Trunk fat mass (kg)** | 14.06 ± 6.44 | 16.28 ± 7.99 | +15.8% | <0.0001 |
| **Leg-to-trunk fat ratio** | 0.895 ± 0.263 | 0.814 ± 0.248 | -9.0% | 0.0048 |
| **Gynoid region % fat** | 42.50 ± 5.14 | 42.78 ± 5.34 | +0.7% | 0.0218 |
| **Android region % fat** | 38.53 ± 8.26 | 40.01 ± 9.49 | +3.9% | 0.0025 |
| **Android/gynoid ratio** | 0.904 ± 0.156 | 0.931 ± 0.163 | +2.9% | 0.0489 |

Regarding body composition, women with a history of cancer exhibited significantly higher trunk fat mass (+15.8%, P < .0001) compared to those without cancer. Absolute leg fat mass was also significantly higher (+4.2%, P = .01); however, the distinct phenotype was driven by the disproportionate accumulation of central adiposity relative to peripheral stores, resulting in a 9.0% lower leg-to-trunk fat ratio (P = .0048; **Error! Reference source not found.**).

### Association Between Peripheral Fat Distribution and Cancer Prevalence

Cancer prevalence demonstrated a significant inverse dose-response relationship with peripheral fat distribution. Weighted cancer prevalence declined from 9.61% in the lowest LTR quartile (android pattern) to 4.29% in the highest quartile (lipedema-like phenotype) (P for trend = .0004; Table 2 and **Error! Reference source not found.**C).

**Table 2.** Distribution of cancer cases across quartiles of leg-to-trunk fat ratio (LTR) among women aged 20-59 years from NHANES 2011-2014 (N=2,818). Q1 represents the lowest LTR (more central fat distribution) and Q4 the highest LTR (more peripheral fat distribution, lipedema-like phenotype). Prevalence was calculated as the unweighted proportion of cancer cases within each quartile. LTR = leg fat mass / trunk fat mass from DXA measurements. P for trend = 0.0004 (Cochran-Armitage test).

| Quartile | N | LTR<br>Range | LTR<br>Mean<br>(SD) | Cancer<br>Cases | Raw<br>Prevalence<br>(n/N) | Raw<br>Prevalence<br>(%) | Weighted<br>Prevalence<br>(%) |
| --- | --- | --- | --- | --- | --- | --- | --- |
| Q1 | 705 | 0.221-0.693 | 0.584<br>(0.084) | 49 | 49/705 | 6.95 | 9.61 |
| Q2 | 704 | 0.693-0.837 | 0.765<br>(0.042) | 35 | 35/704 | 4.97 | 5.84 |
| Q3 | 704 | 0.837-1.021 | 0.926<br>(0.052) | 22 | 22/704 | 3.12 | 4.27 |
| Q4 | 705 | 1.021-2.547 | 1.228<br>(0.180) | 24 | 24/705 | 3.4 | 4.29 |

Note: Weighted prevalence estimates differ from raw prevalence due to NHANES complex survey design. Specifically, cancer cases in the first quartile (Q1) carried higher sampling weights, indicating a higher representation in the general US population than raw counts alone would suggest.

Women in the highest quartile had significantly lower odds of cancer compared to the lowest quartile (unadjusted OR 0.47; 95% CI 0.29–0.78; P = .0036). In sensitivity analyses using binary population cutoffs (Table 4 and **Error! Reference source not found.**A), the inverse direction remained consistent (OR 0.67 at P75; OR 0.59 at P80), though statistical significance attenuated at the 75th percentile (P = .079).

In the primary multivariable model adjusted for BMI, race/ethnicity, smoking, and education (Total Effect), each 1-standard deviation increase in LTR was associated with a 20% reduction in the odds of cancer (OR 0.795; 95% CI 0.666–0.948; P = .011; **Error! Reference source not found.**). When age was introduced into the model (Full Model) to assess mediation, the association was attenuated (OR 0.94; P = .52), consistent with the hypothesis that age captures a substantial portion of the causal pathway linking adipose redistribution to cancer risk (Table 3, **Error! Reference source not found.**).

**Table 3.** Sequential Multivariable Models for the Association Between Leg-to-Trunk Fat Ratio and Cancer Prevalence.

| **Model** | **N** | **Events** | **OR** | **95%**<br>**CI** | **P-value** |
| --- | --- | --- | --- | --- | --- |
| **Unadjusted** | 2818 | 130 | 0.722 | 0.617-0.844 | <0.0001 |
| **Adjusted for BMI** | 2818 | 130 | 0.761 | 0.642-0.903 | 0.0017 |
| **Adjusted for BMI + Race** | 2818 | 130 | 0.769 | 0.646-0.915 | 0.0031 |
| **Adjusted for BMI + Race +**<br>**Smoking** | 2818 | 130 | 0.811 | 0.681-0.965 | 0.0184 |
| **Adjusted for BMI + Race +**<br>**Smoking + Education** | 2818 | 130 | 0.795 | 0.666-0.948 | 0.0109 |
| **Full model (includes age)** | 2818 | 130 | 0.944 | 0.790-1.127 | 0.5216 |

**Table 4.** Sensitivity analysis of the association between the lipedema-like phenotype and cancer prevalence using multiple population prevalence cutoffs. Values represent weighted population prevalence, not raw sample proportions.

| **Cutoff** | **N**<br>**Lipedema**<br>**Phenotype** | **Cancer**<br>**Cases** | **Prevalence**<br>**No**<br>**Lipedema**<br>**(%)** | **Prevalence**<br>**Lipedema**<br>**(%)** | **Odds**<br>**Ratio** | **P-Value** |
| --- | --- | --- | --- | --- | --- | --- |
| **P75** | 705 | 24 | 6.5 | 4.29 | 0.67 | 0.079 |
| **P80** | 564 | 17 | 6.57 | 3.6 | 0.59 | 0.043 |
| **P85** | 423 | 15 | 6.33 | 3.76 | 0.73 | 0.314 |
| **P90** | 282 | 10 | 6.27 | 3.07 | 0.74 | 0.454 |
| **P95** | 141 | 7 | 6.07 | 3.21 | 1.08 | 0.836 |

### Subgroup Analyses and Effect Modification

Stratified analyses revealed that the protective association was most pronounced among women without obesity (BMI < 30 kg/m²). In this group, each 1-SD increase in leg-to-trunk ratio was associated with 33% lower odds of cancer (OR 0.67; 95% CI 0.53–0.85; P = .0007). Among women with obesity, the association followed a similar direction but did not reach statistical significance (OR 0.74; P = .07). The interaction between LTR and obesity status was not statistically significant (P for interaction = .58; Table 5), indicating that the protective direction of the phenotype is consistent regardless of total adiposity.

**Table 5.** Interaction analysis assessing the modifying effects of age, obesity, and ethnicity on the association between the lipedema-like phenotype and cancer.

| Effect Modifier | P-interaction |
| --- | --- |
| Age | 0.2821 |
| Obesity (BMI≥30) | 0.5814 |
| Race/Ethnicity | 0.0741 |
| Smoking Status | 0.7702 |
| Education | 0.8167 |

### Exploratory Analysis by Menopausal Status

To test the potential estrogen-dependence of the phenotype, we stratified the cohort. Among pre-menopausal women, the lipedema-like phenotype (highest LTR quartile) was associated with a 46% lower relative cancer prevalence compared to controls (1.7% vs 3.2%; OR 0.54; 95% CI 0.27–1.08; P = .095). In contrast, among post-menopausal women, this protective association was absent and trended in the opposite direction (10.8% vs 8.5%; OR 1.30; 95% CI 0.70–2.40; P = .403). Although the interaction term did not reach statistical significance (P for interaction = 0.113), the diverging point estimates suggest the protective phenotype is most relevant during the reproductive years (Table S2 and Error! Reference source not found.).

### Discriminative Capacity and Robustness

In model comparison analyses, trunk fat mass provided the strongest improvement in cancer discrimination beyond age (ΔAUC +0.019; P < .001). Both body mass index (ΔAUC +0.0085; P = .023) and the leg-to-trunk ratio (ΔAUC +0.0017; P = .044) yielded statistically significant, albeit modest, gains in discriminative capacity (Table 6 and **Error! Reference source not found.**B).

**Table 6.** Comparison of discriminative capacity (AUC) and model improvement (Likelihood Ratio Test) across different anthropometric indices for cancer prediction.

| Index | AUC (age only) | AUC (age+index) | Delta AUC | LR p-value |
| --- | --- | --- | --- | --- |
| **Leg-Trunk Ratio** | 0.682 | 0.683 | 0.0017 | 0.0435 |
| **BMI** | 0.682 | 0.69 | 0.0085 | 0.0234 |
| **Android/Gynoid Ratio** | 0.682 | 0.683 | 0.0011 | 0.2536 |
| **Gynoid % Fat** | 0.682 | 0.684 | 0.0022 | 0.5385 |
| **Trunk Fat (kg)** | 0.682 | 0.701 | 0.0192 | 0.0004 |

Finally, sensitivity analysis for unmeasured confounding yielded an E-value of 1.83 for the overall estimate and 2.34 for women without obesity. This indicates that an unmeasured confounder would require a risk ratio of at least 2.34 with both the exposure and the outcome to fully explain away the observed protective association in women without obesity, suggesting moderate-to-high robustness of the findings (Table S3 and **Error! Reference source not found.**D).

## DISCUSSION

### Principal Findings

In this cross-sectional analysis of a nationally representative sample of US women, a lipedema-like peripheral fat distribution was inversely associated with cancer prevalence. The robustness of this linear trend (P < .001) suggests that the immunometabolic benefit of peripheral adiposity follows a biological continuum rather than a pathological threshold (11–13). Critically, this protective effect was most robust among women without obesity (OR 0.67; P = .0007). Among women with obesity, a similar inverse trend was observed (OR 0.74; P = .07), and the lack of significant interaction (P = .58) indicates that the distinct adipose distribution remains a relevant factor independent of total adiposity.

Although adjusting for age attenuated the primary association in the full model, this is expected given the strong collinearity between aging and peripheral fat loss (14, 15). Importantly, stratified analyses demonstrated that women with cancer exhibited lower LTR than controls within every pre-menopausal age decade (Table S1 and **Error! Reference source not found.**). The persistence of this difference specifically during the reproductive years argues that the phenotype is intrinsic to body composition rather than a statistical artifact of aging. Collectively, these data support the "metabolic sink" hypothesis, suggesting the phenotype serves as a marker of metabolic health that remains relevant even in the absence of overt caloric excess (11).

### Comparison With Prior Literature

Our results extend previous work distinguishing harmful central adiposity from protective peripheral depots. The European Prospective Investigation into Cancer and Nutrition reported that waist circumference and waist-to-hip ratio predicted cancer mortality independently of BMI, with hazard ratios of 1.17 and 1.25 per standard deviation increment, respectively(16). Conversely, gluteofemoral adiposity has been linked to favorable metabolic profiles, including lower inflammatory markers and improved insulin sensitivity (17–22). In our analysis, trunk fat mass remained the strongest predictor of cancer risk (ΔAUC 0.019; P < .001), while the leg-to-trunk ratio showed an inverse trend. These data reinforce central adiposity as the principal driver of risk, while suggesting that a lipedema-like pattern may attenuate the carcinogenic burden associated with obesity.

### Biological Mechanisms: The Metabolic Sink and Menopausal Switch

Our findings align with the "metabolic sink" hypothesis, wherein gluteofemoral adipose tissue buffers circulating lipids, preventing ectopic deposition and lipotoxicity. Unlike visceral fat, which releases pro-inflammatory cytokines into the portal circulation, gluteofemoral fat exhibits lower lipolytic activity (11, 23–25). and higher secretion of adiponectin—an adipokine with established anti-tumor properties, including suppression of proliferation via AMPK activation (26–28), inhibition of angiogenesis (29–31), and attenuation of the pro-inflammatory tumor microenvironment through NF-κB suppression (27).

This protective capacity appears estrogen-dependent. Our exploratory analysis revealed a "menopausal switch": the inverse association was observed in premenopausal women (OR 0.54) but was absent in the postmenopausal state. This aligns with physiological evidence that estrogen withdrawal compromises the metabolic buffering function of peripheral fat, potentially shifting it from a protective depot to a dysfunctional one (32–39).

### The Immunological Shield

Beyond metabolism, the lipedema-like phenotype may confer an immunological advantage. Previous analyses of this cohort demonstrated that women with lipedema-like fat distribution exhibit 44% lower insulin resistance, 7.6% lower neutrophil-to-lymphocyte ratio, and a striking 79% reduction in diabetes odds (40–42).

Complementing these findings, we recently characterized a paradoxical immunoglobulin signature in women with clinical lipedema: more frequent food-specific IgG positives despite markedly lower total IgG production—a pattern consistent with mucosal barrier dysfunction coupled with systemic downregulation of humoral immunity (7). The consistency of protective associations across metabolic (diabetes), inflammatory (NLR), immunological (IgG dysregulation), autoimmune (celiac disease, thyroid disease), and now neoplastic outcomes supports the concept of an integrated immunometabolic shield conferred by peripheral adiposity.

This creates a fascinating genotype-phenotype paradox. Despite a known high prevalence of autoimmune risk alleles (e.g., HLA-DQ2/DQ8) in lipedema patients, the phenotypic expression of disease appears suppressed (8). The expanded gluteofemoral depot may create a systemic environment that raises the threshold for both autoimmune activation and malignant transformation, effectively decoupling genetic susceptibility from clinical disease.

### Clinical Implications

These findings challenge the prevailing paradigm that treats obesity as a uniform cancer risk factor (43, 44). BMI, which conflates central and peripheral adiposity, may inadequately stratify cancer risk (45–47). Interventions focused solely on weight reduction may be misguided for women with predominantly peripheral fat distribution, in whom caloric restriction could paradoxically reduce protective adipose reserves without addressing the visceral fat that drives metabolic dysfunction (48–54).

More critically, the identification of gluteofemoral tissue as a potential immunometabolic buffer raises questions regarding large-volume liposuction for lipedema. If this depot functions as an active endocrine organ buffering inflammation and potentially suppressing carcinogenesis, aggressive surgical removal could disrupt this protective equilibrium (48–50, 54, 55). We hypothesize that rapid excision could inadvertently alter the cancer risk trajectory in susceptible individuals or force future lipid deposition into the visceral compartment—since our findings indicate that cancer risk is driven by the trunk-to-leg disproportion rather than leg adiposity itself (56, 57).

We do not advocate abandoning surgical treatment, which can provide symptomatic relief(58). Rather, we call for a paradigm shift: lipedema tissue should be conceptualized not merely as diseased fat to be excised, but as a functional organ whose systemic contributions must be weighed against local symptoms.

### Evolutionary Perspective

From an evolutionary standpoint, the lipedema-like phenotype may represent an ancestral adaptation for energy conservation that prioritizes tissue preservation over hyper-vigilant inflammatory responses (59). The capacity to store energy safely in the lower extremities—without the metabolic penalties of visceral accumulation—would confer survival advantages during periods of food scarcity, particularly during pregnancy and lactation (60). The inverse associations with diabetes, autoimmunity, and now cancer suggest this "thrifty phenotype" is coupled with a calibrated immune system (61). Thus, lipedema may not be a disease in the classical sense, but rather an extreme expression of a protective evolutionary strategy—with both advantages (metabolic and oncological protection) and disadvantages (adipose expansion) that must be balanced in clinical decision-making (62, 63).

### Limitations

Several limitations merit consideration. First, the cross-sectional design precludes causal inference; observed associations represent cancer prevalence (survivorship) rather than incidence. Cancer diagnoses were self-reported, and some malignancies likely preceded body composition measurement, raising the possibility of reverse causation (64). However, the persistence of the protective signal among women with obesity—who typically maintain adipose reserves—and the exclusion of underweight individuals mitigates the likelihood that cachexia alone explains these findings.

Second, DXA-derived LTR is a proxy that cannot distinguish clinically diagnosed lipedema from other forms of peripheral obesity, nor capture the pain, edema, or fibrosis central to the syndrome (1, 65).

Third, residual confounding by unmeasured factors (physical activity, diet, reproductive history, medication use) may have influenced associations; however, the E-value of 2.34 for the subgroup without obesity indicates that an unmeasured confounder would require a strong association with both exposure and outcome to fully explain the findings (66).

Finally, site-specific cancer numbers were insufficient to examine whether protection is confined to particular malignancies linked to adiposity and sex steroid metabolism. While we excluded age from primary models to capture the total effect (viewing age as a mediator per DAG theory), we provided adjusted models for transparency (67).

### Future Directions

Longitudinal cohorts with repeated DXA measures and adjudicated cancer outcomes are needed to determine if changes in regional adiposity predict incident malignancy. Mechanistic studies should characterize the adipokine, cytokine, and immune cell profiles of lipedema tissue to identify molecular correlates of this "shield."(68) Most urgently, prospective registries of women undergoing lipedema liposuction should monitor for new autoimmune or neoplastic events to quantify trade-offs between symptom relief and loss of a potentially protective depot. Until such data are available, clinicians should approach lipedema with appreciation for its potential systemic protective functions(69, 70).

## CONCLUSIONS

In this nationally representative study of US women, peripheral fat distribution characteristic of the lipedema-like phenotype was inversely associated with cancer prevalence in a significant dose-response manner (P for trend = .0004), with the strongest association observed among women without obesity and a consistent directional trend among women with obesity. When integrated with prior evidence of favorable metabolic, inflammatory, and autoimmune profiles in women with disproportionate gluteofemoral adiposity, these findings support reframing lipedema from a purely pathologic condition toward a complex immunometabolic phenotype that may confer partial protection against the systemic consequences of obesity. This paradigm shift should inform risk stratification, therapeutic decision-making, and future research addressing the long-term implications of modifying regional adipose depots.

## FIGURE LEGENDS

**Figure 1.**
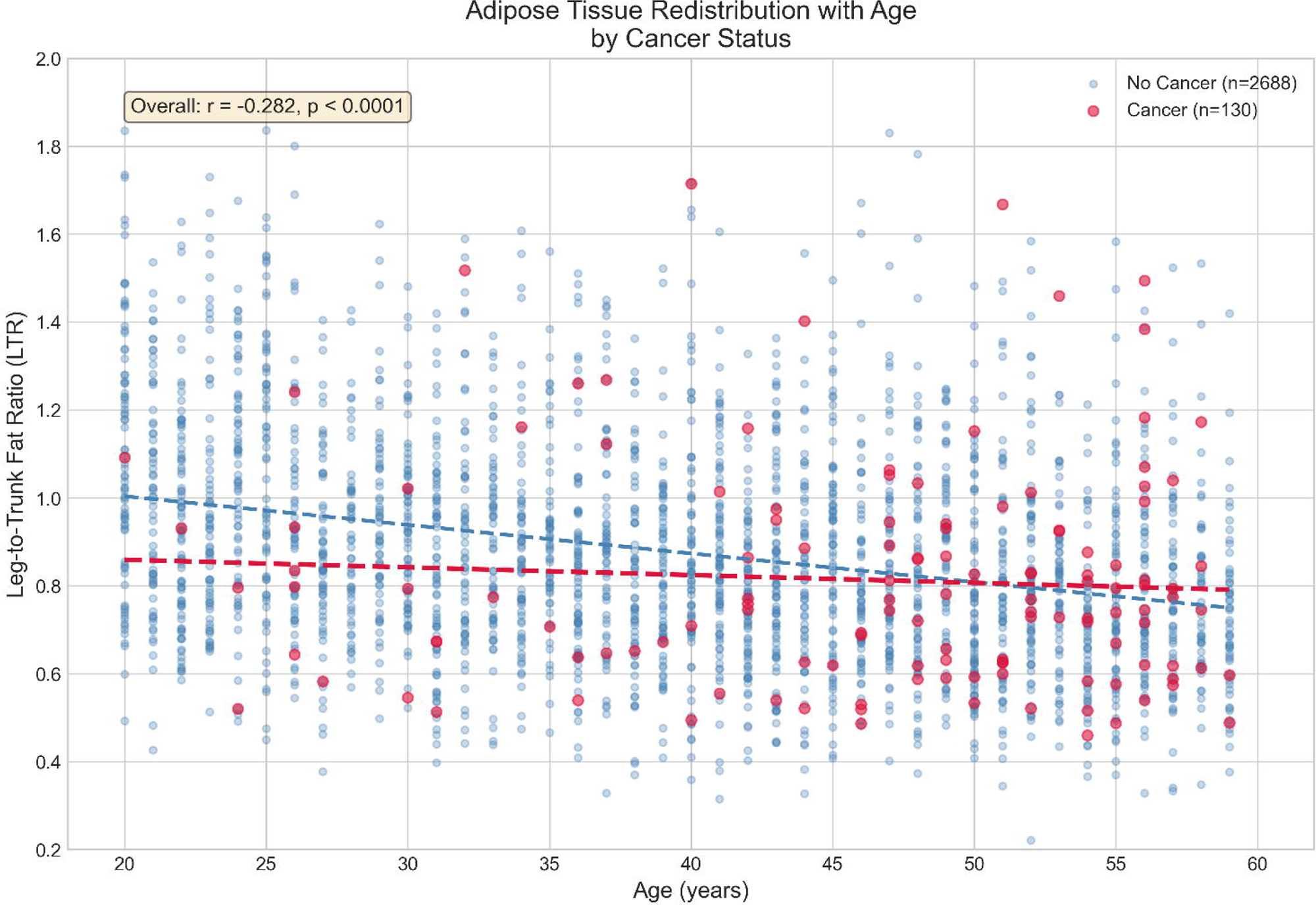
Distribution of anthropometric and body composition variables stratified by cancer diagnosis. Women with a history of cancer exhibit a distinct phenotype characterized by significantly higher trunk fat mass and a lower leg-to-trunk fat ratio compared to those without cancer, despite similar overall BMI distributions in many cases.

**Figure 2.**
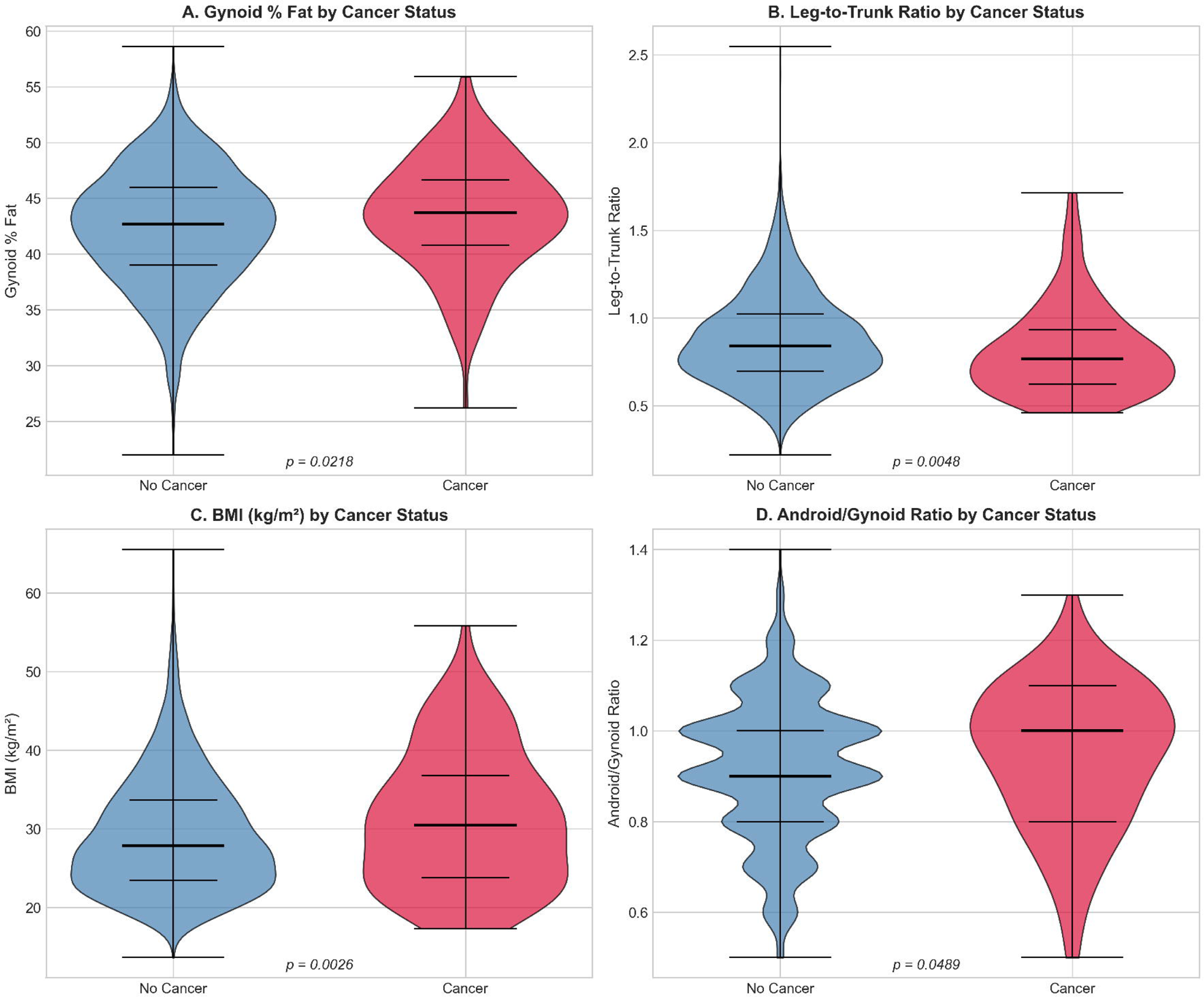
Age-related redistribution of adipose tissue stratified by cancer history. The scatter plot demonstrates the physiological decline in Leg-to-Trunk Fat Ratio (LTR) associated with aging (blue dashed line; Overall r = -0.282, p < 0.0001). Notably, women with a history of cancer (red line, n=130) exhibit a distinct phenotype characterized by lower peripheral adiposity relative to controls (blue line, n=2,688), particularly during the pre-menopausal years (ages 20–50). The regression lines converge in the post-menopausal period (∼55–60 years), supporting the hypothesis that the protective "metabolic sink" function of peripheral fat may be estrogen-dependent.

**Figure 3.**
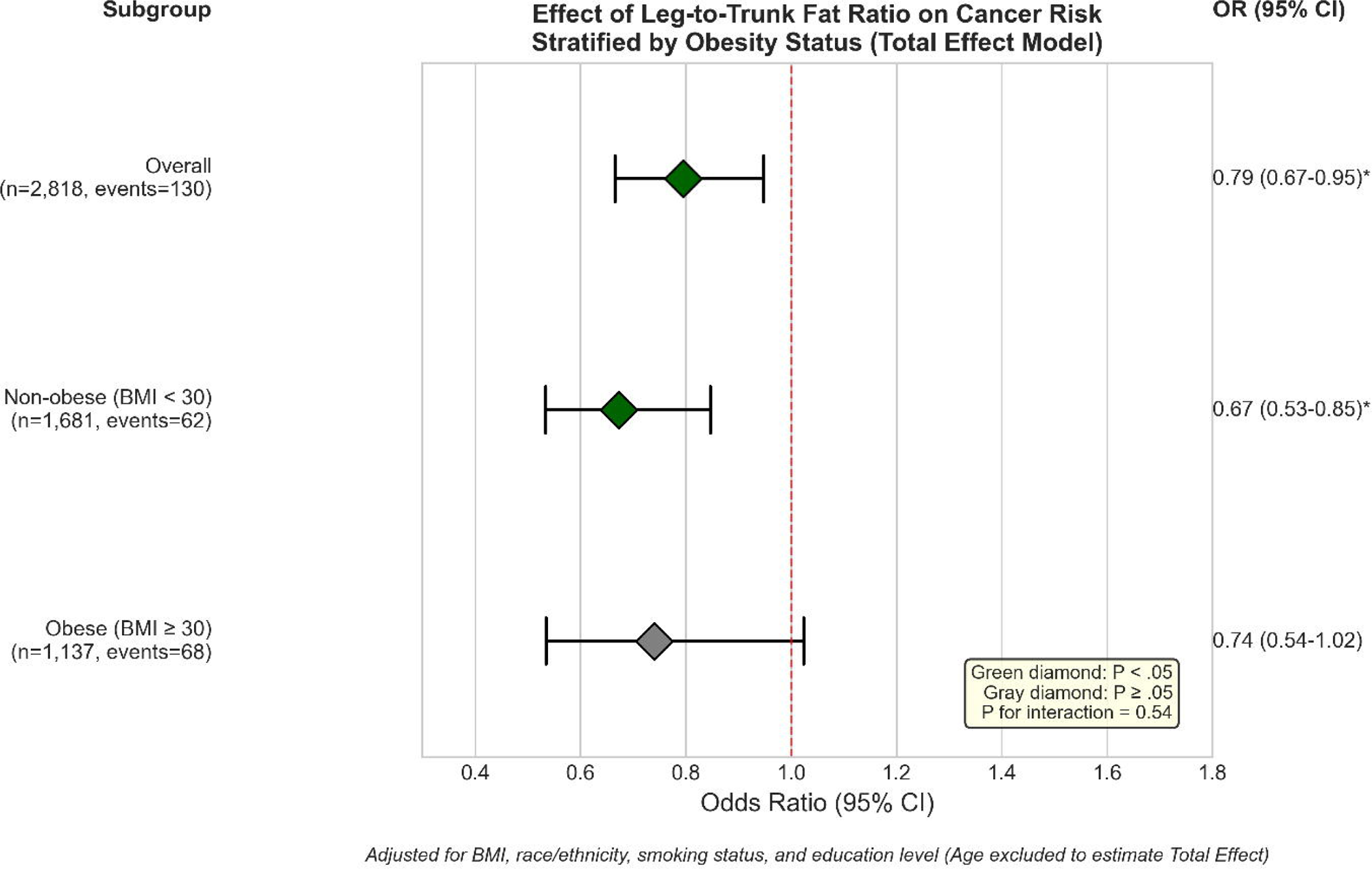
Forest plot of multivariable-adjusted odds ratios (95% CI) for cancer prevalence associated with leg-to-trunk fat ratio (LTR), stratified by obesity status. Odds ratios represent the change in cancer odds per 1-SD increase in LTR, adjusted for BMI, race/ethnicity, smoking status, and education level (age excluded to estimate total effect). Green diamonds indicate statistically significant associations (P < .05); the gray diamond indicates a non-significant trend (P ≥ .05). The protective association was robust in women without obesity (OR 0.67; 95% CI 0.54–0.85) and showed a consistent directional trend among women with obesity (OR 0.74; 95% CI 0.54–1.03), with no significant effect modification by obesity status (P for interaction = 0.54).

**Figure 4.**
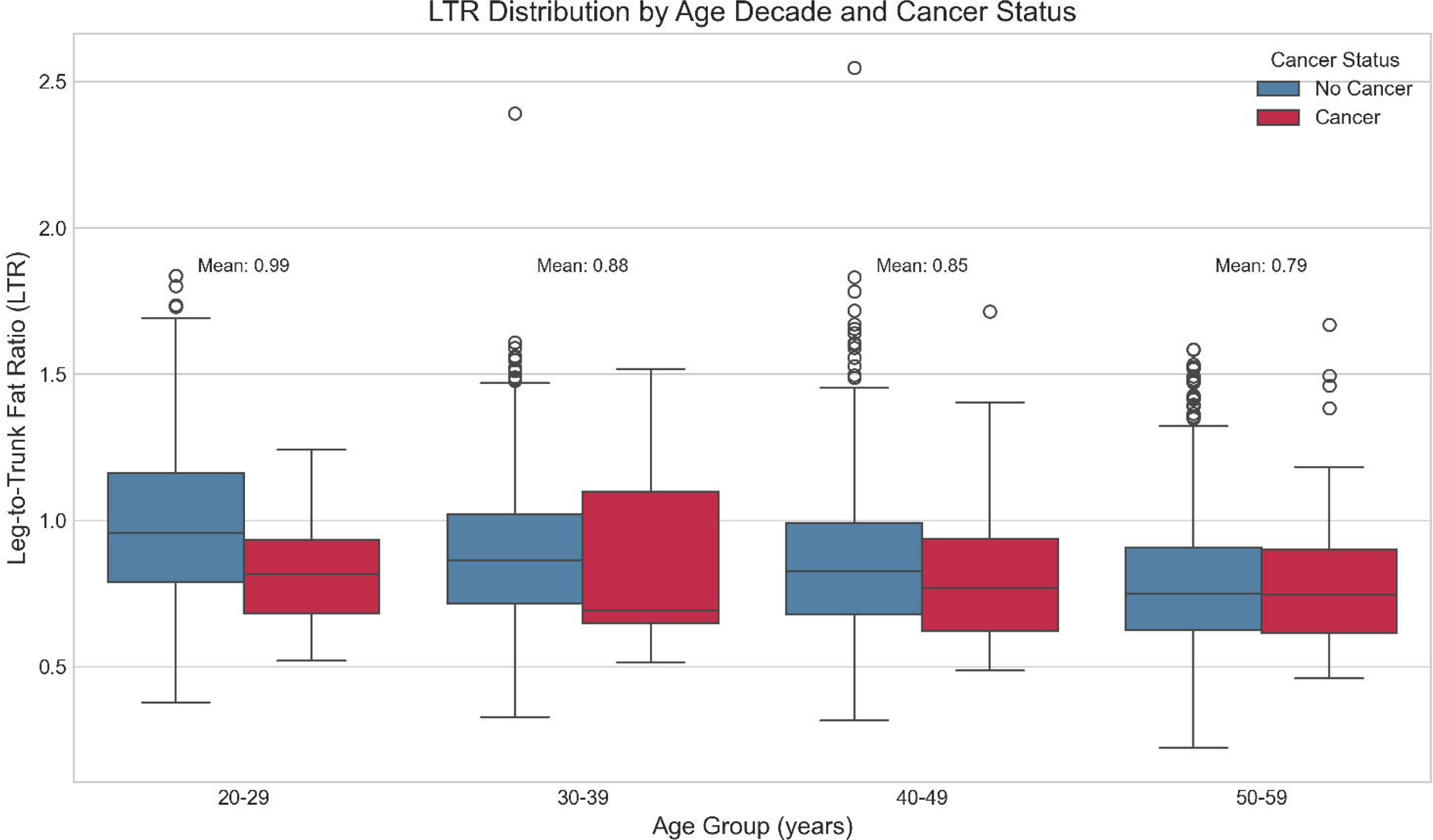
Consistency of the inverse association across the lifespan. Women with a history of cancer exhibit lower Leg-to-Trunk Ratios compared to controls within every decade of life, demonstrating that the phenotype persists independent of aging.

**Figure 5.**
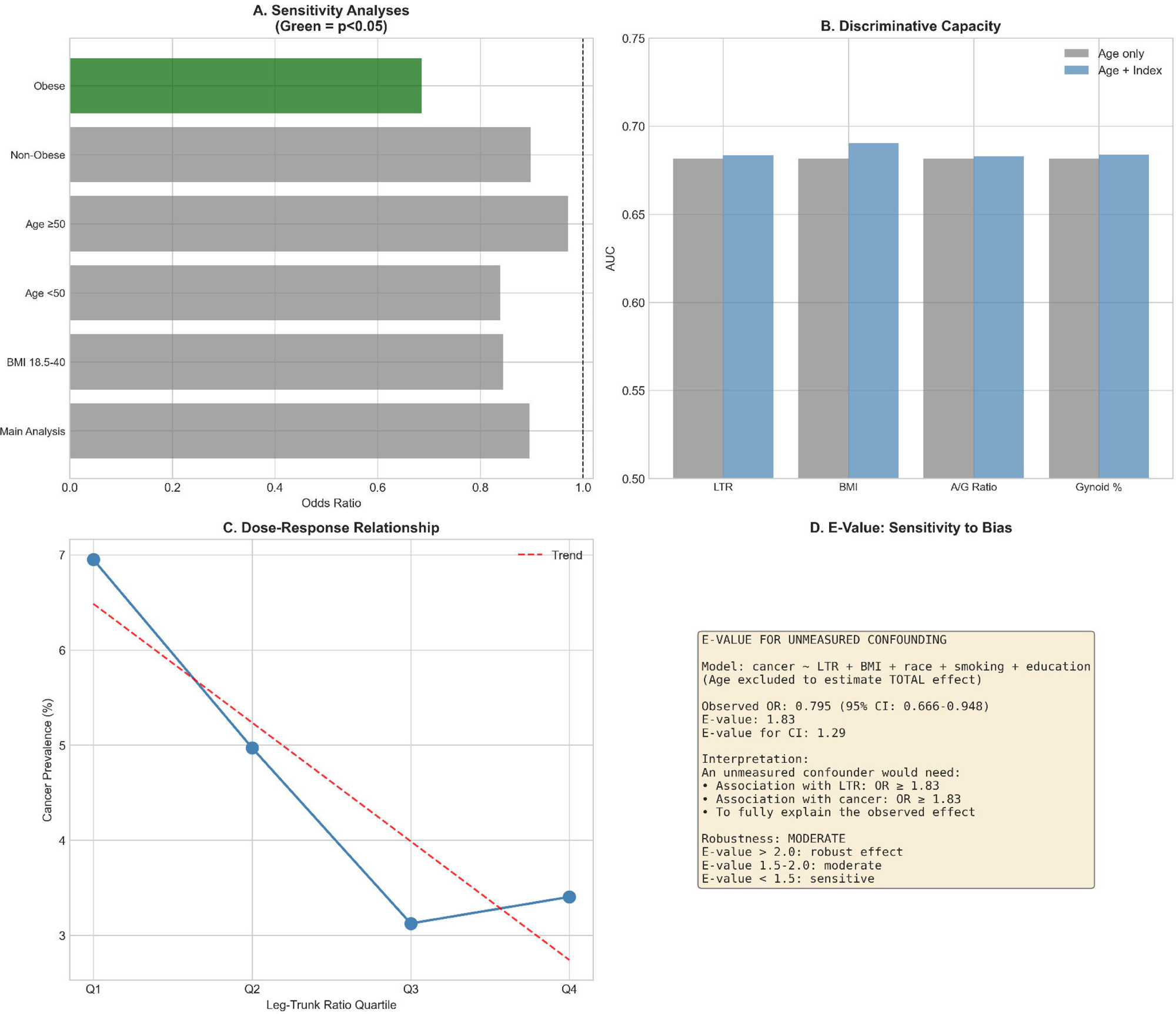
Sensitivity, Discrimination, and Robustness Analyses of the Association Between Leg-to-Trunk Fat Ratio (LTR) and Cancer. (A) Sensitivity Analyses: Odds ratios (OR) per 1-SD increase in LTR across subgroups (adjusted for age and BMI). Red bars indicate statistical significance (P < 0.05). (B) Discriminative Capacity: Comparison of AUC for prediction models using age alone (gray) versus age plus anthropometric index (blue). (C) Dose-Response: Cancer prevalence across LTR quartiles (Q1=android to Q4=lipedema-like phenotype). Red dashed line indicates linear trend (P trend = 0.0004). (D) E-Value: Analysis quantifying the minimum strength of unmeasured confounding required to explain away the observed protective effect.

**Figure 6.**
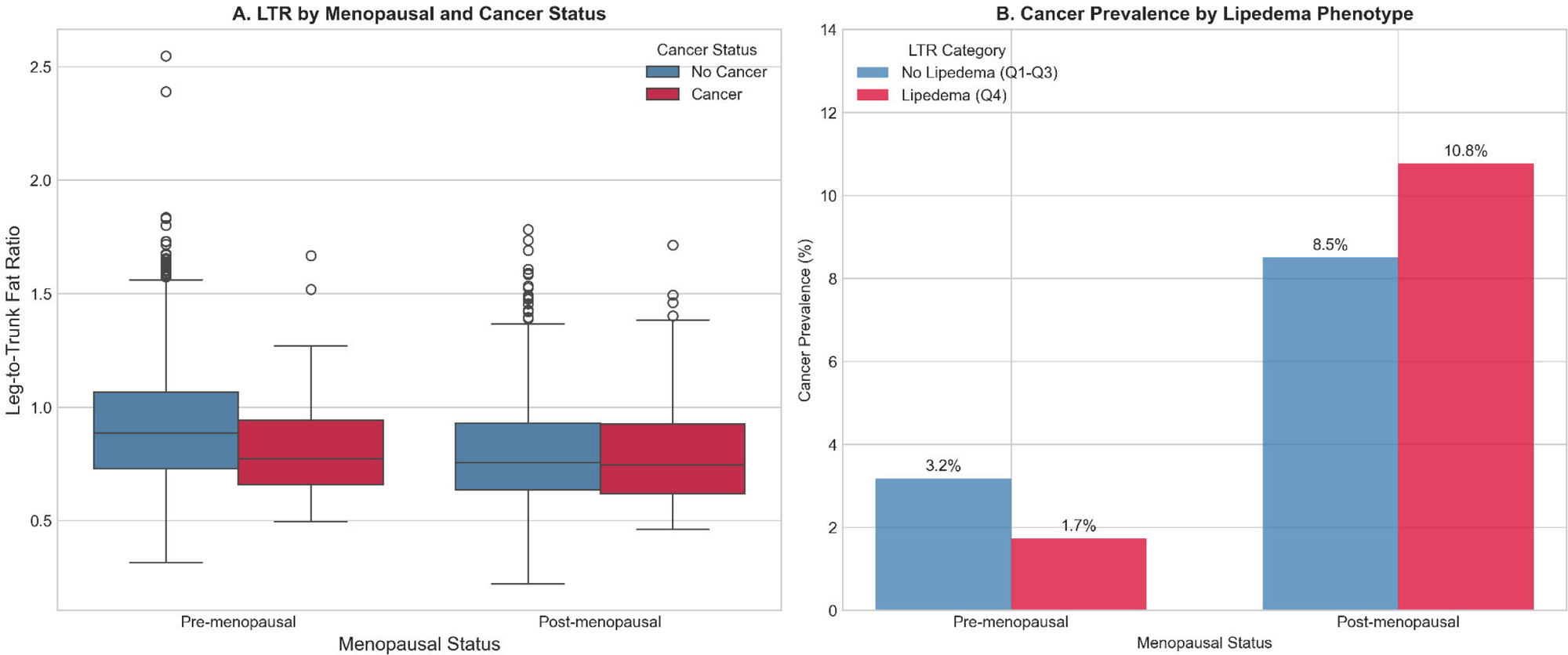
The "Menopausal Switch": Potential Estrogen-Dependent Modulation of the Lipedema Phenotype-Cancer Association. (A) Distribution of leg-to-trunk fat ratio (LTR) stratified by menopausal and cancer status. (B) Cancer prevalence stratified by menopausal status and lipedema phenotype. Among pre-menopausal women, the lipedema phenotype shows an inverse association with cancer (3.2% vs 1.7% prevalence; OR = 0.54). In post-menopausal women, this pattern is distinct, with higher prevalence observed in the lipedema phenotype group compared to controls (10.8% vs 8.5%; OR = 1.30). This contrast aligns with the hypothesis that the metabolic sink function of gluteofemoral adipose tissue may be estrogen dependent. Note: Lipedema phenotype is defined as LTR ≥ 75th percentile (Q4); No Lipedema is defined as LTR < 75th percentile (Q1–Q3).

## DATA AVAILABILITY STATEMENT

The data that support the findings of this study are openly available from the National Center for Health Statistics (NCHS) at https://www.cdc.gov/nchs/nhanes/.

## ACKNOWLEDGMENTS

We utilized Claude Opus 4.1 and GEMINI 3 Pro to assist in the generation and debugging of the Python code used for statistical analysis, specifically to ensure the correct implementation of NHANES complex survey design weights.

