## Supplement for "Lipedema-like Phenotype and Cancer Prevalence in US Women: A Cross-Sectional Analysis of NHANES 2011–2014"

**Title:** Lipedema-like Phenotype and Cancer Prevalence in US Women: A Cross-Sectional Analysis of NHANES 2011–2014

**Authors:** Alexandre C. M. Amato, Juliana L. S. Amato, Daniel A. Benitti

**Contents:**

- **Figure S1:** Participant Flow Chart
- **Table S1:** Age-stratified comparison of Leg-to-Trunk Fat Ratio
- **Table S2:** Association stratified by Menopausal Status
- **Table S3:** Sensitivity analysis (E-values)

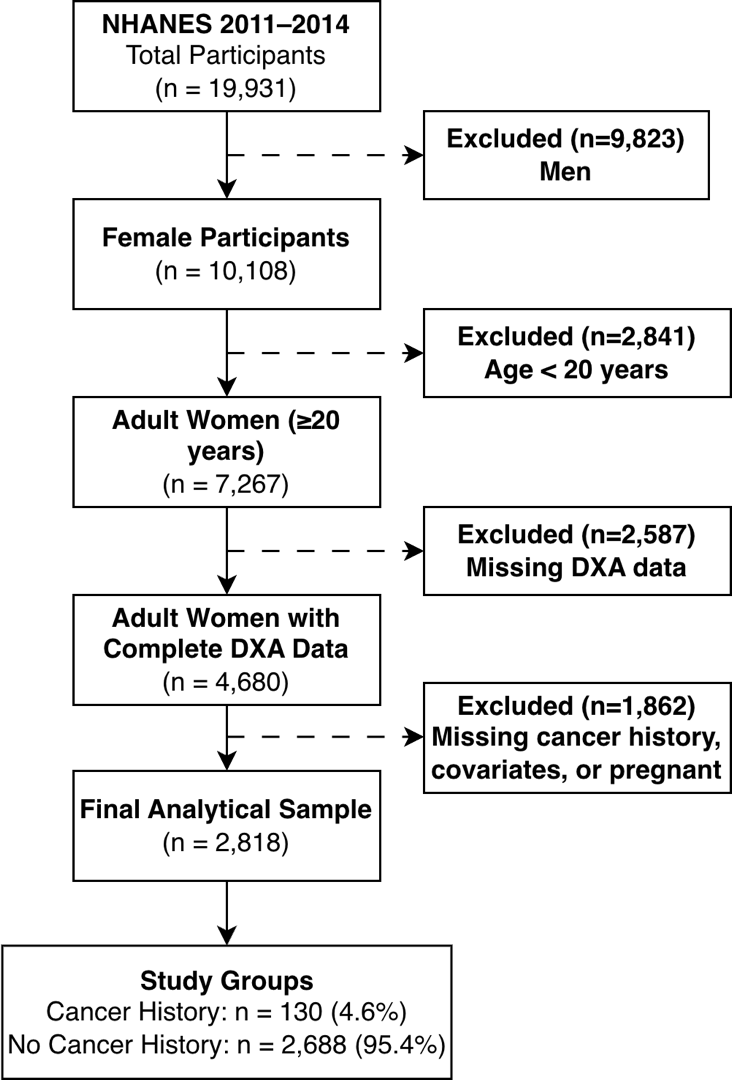

**Figure S1. Participant Flow Chart.** STROBE-compliant flow diagram illustrating participant selection from NHANES 2011–2014. From the initial 19,931 survey participants, sequential exclusion of men (n=9,823), participants aged <20 years (n=2,841), those with missing DXA data (n=2,587), and those with missing cancer history, covariates, or current pregnancy (n=1,862) yielded a final analytical sample of 2,818 adult women. Of these, 130 (4.6%) reported a prior cancer diagnosis.

**Table S1** - Age-stratified comparison of Leg-to-Trunk Fat Ratio (LTR) between women with and without cancer. Data are presented as weighted mean ± standard deviation. The "Difference" column reflects the relative percent difference in the cancer group compared to controls. P-values derived from design-adjusted t-tests.

| **Age Decade** | **N** | **Cancer Cases** | **Cancer Prev (%)** | **LTR No Cancer** | **LTR Cancer** | **Difference (%)** | **P-value** |
| --- | --- | --- | --- | --- | --- | --- | --- |
| **20-29** | 681 | 10 | 1.5 | 0.989 ± 0.268 | 0.837 ± 0.224 | -15.4% | 0.0753 |
| **30-39** | 687 | 18 | 2.6 | 0.885 ± 0.244 | 0.844 ± 0.301 | -4.7% | 0.4762 |
| **40-49** | 752 | 43 | 5.7 | 0.853 ± 0.252 | 0.804 ± 0.246 | -5.7% | 0.2209 |
| **50-59** | 698 | 59 | 8.5 | 0.785 ± 0.228 | 0.806 ± 0.260 | +2.7% | 0.4972 |

***Note****: Women with a history of cancer exhibited lower LTR in all pre-menopausal age decades (20–49 years), consistent with the lipedema-like phenotype. In the 50–59 age stratum, this pattern is attenuated or reversed, aligning with the "menopausal switch" hypothesis described in the main text.*

**Table S2.** Association Between Lipedema-like Phenotype and Cancer Prevalence Stratified by Menopausal Status.

| **Menopausal Status** | **LTR Category** | **Total N** | **Cancer Cases** | **Prevalence (%)** | **OR (95% CI)** | **P-value** |
| --- | --- | --- | --- | --- | --- | --- |
| **Pre-menopausal** | No Lipedema (Q1-Q3) | 1384 | 44 | 3.2 | Reference | - |
| **Pre-menopausal** | Lipedema (Q4) | 575 | 10 | 1.7 | 0.54 (0.27-1.08) | 0.095 |
| **Post-menopausal** | No Lipedema (Q1-Q3) | 729 | 62 | 8.5 | Reference | - |
| **Post-menopausal** | Lipedema (Q4) | 130 | 14 | 10.8 | 1.30 (0.70-2.40) | 0.403 |

***Note:*** *P for interaction (lipedema category × menopausal status) = 0.113 .*

**Table S3.** Sensitivity analysis for unmeasured confounding using E-values.

| **Subgroup** | **N** | **Cancer Cases** | **OR** | **CI_Low** | **CI_High** | **P_value** | **E_value** | **E_value_CI** | **Robustness** |
| --- | --- | --- | --- | --- | --- | --- | --- | --- | --- |
| **Overall** | 2818 | 130 | 0.795 | 0.666 | 0.948 | 0.0109 | 1.83 | 1.29 | MODERATE |
| **Non-obese (BMI<30)** | 1681 | 62 | 0.673 | 0.534 | 0.847 | 0.0007 | 2.34 | 1.64 | ROBUST |
| **Obese (BMI>=30)** | 1137 | 68 | 0.741 | 0.535 | 1.024 | 0.0697 | 2.04 | 1.18 | ROBUST |
| **Age <50** | 2120 | 71 | 0.817 | 0.633 | 1.055 | 0.1215 | 1.75 | 1.3 | MODERATE |
| **Age >=50** | 698 | 59 | 1.011 | 0.784 | 1.303 | 0.9355 | 1.11 | 1.87 | SENSITIVE |

***Note****: The E-value quantifies the minimum strength of association that an unmeasured confounder would require with both the exposure (leg-to-trunk fat ratio) and the outcome (cancer) to fully explain away the observed effect. Higher values indicate greater robustness; for instance, the E-value of 2.34 in non-obese women implies that an unmeasured confounder would need to increase risk by at least 2.34-fold to nullify the association.*
